# Deep Learning Estimation of Small Airways Disease from Inspiratory Chest CT is Associated with FEV_1_ Decline in COPD

**DOI:** 10.1101/2024.09.10.24313079

**Authors:** Muhammad F. A. Chaudhary, Hira A. Awan, Sarah E. Gerard, Sandeep Bodduluri, Alejandro P. Comellas, Igor Z. Barjaktarevic, R. Graham Barr, Christopher B. Cooper, Craig J. Galban, MeiLan K. Han, Jeffrey L. Curtis, Nadia N. Hansel, Jerry A. Krishnan, Martha G. Menchaca, Fernando J. Martinez, Jill Ohar, Luis G. Vargas Buonfiglio, Robert Paine, Surya P. Bhatt, Eric A. Hoffman, Joseph M. Reinhardt

## Abstract

**Rationale:** Quantifying functional small airways disease (fSAD) requires additional expiratory computed tomography (CT) scan, limiting clinical applicability. Artificial intelligence (AI) could enable fSAD quantification from chest CT scan at total lung capacity (TLC) alone (fSAD^TLC^).

**Objectives:** To evaluate an AI model for estimating fSAD^TLC^ and study its clinical associations in chronic obstructive pulmonary disease (COPD).

**Methods:** We analyzed 2513 participants from the SubPopulations and InteRmediate Outcome Measures in COPD Study (SPIROMICS). Using a subset (*n* = 1055), we developed a generative model to produce virtual expiratory CTs for estimating fSAD^TLC^ in the remaining 1458 SPIROMICS participants. We compared fSAD^TLC^ with dual volume, parametric response mapping fSAD^PRM^. We investigated univariate and multivariable associations of fSAD^TLC^ with FEV_1_, FEV_1_/FVC, six-minute walk distance (6MWD), St. George’s Respiratory Questionnaire (SGRQ), and FEV_1_ decline. The results were validated in a subset (*n* = 458) from COPDGene study. Multivariable models were adjusted for age, race, sex, BMI, baseline FEV_1_, smoking pack years, smoking status, and percent emphysema.

**Measurements and Main Results:** Inspiratory fSAD^TLC^ was highly correlated with fSAD^PRM^ in SPIROMICS (Pearson’s R = 0.895) and COPDGene (R = 0.897) cohorts. In SPIROMICS, fSAD^TLC^ was associated with FEV_1_ (L) (adj.β = −0.034, *P* < 0.001), FEV_1_/FVC (adj.β = −0.008, *P* < 0.001), SGRQ (adj.β = 0.243, *P* < 0.001), and FEV_1_ decline (mL / year) (adj.β = −1.156, *P* < 0.001). fSAD^TLC^ was also associated with FEV_1_ (L) (adj.β = −0.032, *P* < 0.001), FEV_1_/FVC (adj.β = −0.007, *P* < 0.001), SGRQ (adj.β = 0.190, *P* = 0.02), and FEV_1_ decline (mL / year) (adj.β = - 0.866, *P* = 0.001) in COPDGene. We found fSAD^TLC^ to be more repeatable than fSAD^PRM^ with intraclass correlation of 0.99 (95% CI: 0.98, 0.99) vs. 0.83 (95% CI: 0.76, 0.88).

**Conclusions:** Inspiratory fSAD^TLC^ captures small airways disease as reliably as fSAD^PRM^ and is associated with FEV_1_ decline.

**Funding Source:** This work was supported by NHLBI grants R01 HL142625, U01 HL089897 and U01 HL089856, by NIH contract 75N92023D00011, and by a grant from The Roy J. Carver Charitable Trust (19-5154). The COPDGene study (NCT00608764) has also been supported by the COPD Foundation through contributions made to an Industry Advisory Committee that has included AstraZeneca, Bayer Pharmaceuticals, Boehringer-Ingelheim, Genentech, GlaxoSmithKline, Novartis, Pfizer, and Sunovion.

## INTRODUCTION

Small conducting airways of the lungs are the primary sites of airflow limitation in early obstructive lung disease (1, 2), and their loss is extensive by the time that airflow limitation is detectable by spirometry (3). Identifying early small airways disease (SAD) in susceptible individuals who have inhalational exposures linked to COPD is essential to the development of disease-modifying therapies for this leading cause of worldwide death and disability.

Currently, the only *in vivo* method to quantify SAD relies on image registration for demonstrating non-emphysematous air-trapping on chest computed tomography (CT) scans obtained at inspiration and expiration (4). These co-registered scans are analyzed through parametric response mapping (PRM) to quantify such air-trapping, termed functional small airways disease (fSAD^PRM^) (4). MicroCT analysis of resected lungs demonstrated that fSAD^PRM^ accurately reflects SAD in advanced COPD (5). However, the need for additional expiratory CT incurs increased cost and ionizing radiation exposure. Additionally, expiratory CT scan acquisition in clinical settings requires specialized technician training and there is no harmonized protocol to coach and acquire images at different lung volumes (6). These drawbacks preclude application of PRM analysis to clinical settings where expiratory CT scans are unavailable. By contrast, inspiratory chest CT scans are common in many clinical settings, including lung cancer screening. Hence, a method for estimating fSAD from inspiratory chest CT scans alone would be valuable.

Deep generative modeling can reliably generate multimodal medical images by converting a medical image from one modality to the other (7). Recently, we developed a method for estimating an expiratory chest CT scan solely from a given inspiratory CT image (7). The synthesized virtual expiratory image could be used in combination with the available inspiratory CT scan for estimating SAD. We hypothesize that this single inspiratory volume fSAD estimation from chest CT at total lung capacity (TLC) can be used to identify regions of SAD and that these regions will be associated with poor outcomes in COPD. To test our hypothesis, we analyzed data from the SubPopulations and InteRmediate Outcome Measures in COPD Study (SPIROMICS) cohort (8). The results were validated in a cohort from the Genetic Epidemiology of COPD (COPDGene) study (9). We also investigated the repeatability of inspiratory CT fSAD and compared its repeatability with the conventional fSAD^PRM^ extracted from two volumes.

## METHODS

### Study Populations

We analyzed data from the SubPopulations and InteRmediate Outcome Measures in COPD Study (SPIROMICS) which is a multicenter prospective cohort study being conducted at 14 clinical centers across the United States (US) (8). SPIROMICS enrolled 2981 participants between 40 and 80 years of age across four strata: with smoking history ≤ 1 pack-year (stratum 1) or a smoking history ≥ 20 pack-years (strata 2 – 4). The participants underwent high-resolution chest CT scans on full inspiration and expiration, i.e., at both total lung capacity (TLC) and residual volume (RV), respectively at their enrollment visit. We divided SPIROMICS participants as described below into two non-overlapping subsets, which were respectively used to train our generative model (to transform inspiratory CT to synthetic expiratory CT scans), and for the association of the resulting fSAD measurements from the single inspiratory and synthetic expiratory scans.

We externally validated our model on a subset from the Genetic Epidemiology of COPD (COPDGene) study (9). COPDGene is a multicenter observational cohort study that recruited nearly 10,300 participants across 21 clinical centers in the US. COPDGene used a different image acquisition protocol from SPIROMICS. COPDGene acquired chest CT scans at TLC and functional residual capacity (FRC). However, in a sub-study at a single clinical site (Iowa), some participants were also scanned at RV (*n* = 458), and this group was used in this analysis.

Both studies were conducted according to the principles of the Helsinki accord. Written informed consent for both studies was provided by all participants, and the protocols were approved by the Institutional Review Boards (IRBs) of each participating study center.

### Estimating fSAD from Inspiratory Chest CT

To synthesize virtual expiratory CT scans from a participant’s inspiratory chest CT scan alone, we used a recently developed generative adversarial network (GAN), called LungViT (7). Before training the model, we registered the expiratory chest CT scan (treated as moving image) to the inspiratory chest CT scan (treated as the registration reference). We trained the generative model using registered scan pairs, obtained at TLC and RV, from SPIROMICS. The training set (*n* = 1055) comprised 55 individuals who never smoked, and 200 participants randomly sampled from each COPD Global Initiative for Chronic Lung Disease (GOLD) stage (0–4).

GANs use image pairs to learn a transformation from one image to the other, in our case, to transform TLC scans to RV scans. Generative models rely on two coupled neural networks, where an additional model (called the discriminator) is used to enhance and assess the image quality generated by the generative model (the generator) (7). Iterative feedback from the discriminator helps the generator to produce perceptually realistic images. Once trained, the generator can be used to synthesize a virtual chest CT scan at RV from a given TLC CT scan alone. For more details on model architecture and training please refer to the work in (7) and the online supplement.

Next, we used the trained generator to synthesize virtual expiratory chest CT scans for the remaining SPIROMICS (*n* = 1458) and COPDGene participants (*n* = 458). These virtual expiratory CT scans were employed, in combination with the TLC scans, to compute a variable called fSAD^TLC^. We calculated fSAD^TLC^ as the voxels between −950 HU to −810 HU on inspiratory chest CT scan and between −1000 HU to −857 HU on the virtual or synthetic expiratory CT scan (4, 10). We compared the proposed fSAD^TLC^ with the conventional dual volume, image registration-based fSAD, obtained using the PRM (fSAD^PRM^). The fSAD^PRM^ was computed by registering the expiratory CT scans at RV to the inspiratory CT scans at TLC. The fSAD^PRM^ was defined as voxels between −950 HU and −810 HU on inspiratory chest CT and between −1000 HU and −857 HU on the expiratory CT (4, 10). The only difference between the fSAD^PRM^ and fSAD^TLC^ calculation was that the latter used a virtual expiratory CT scan at RV instead of the actual RV scan acquired by SPIROMICS. Both fSAD^PRM^ and fSAD^TLC^ were expressed as a percentage of total lung volume.

### Predictors

Predictors in this study included age, sex, race, body mass index (BMI), smoking status (current or former defined as ≥ 6-month cessation), smoking pack years, post-bronchodilator forced expiratory volume in 1 second (FEV_1_), percent emphysema defined as percentage of low-attenuation areas (LAA%) below −950 Hounsfield units (HU), and fSAD^TLC^ or fSAD^PRM^.

### Outcomes

We studied the association of fSAD^TLC^ with various functional and clinical outcomes of respiratory morbidity in both SPIROMICS and COPDGene cohorts. Our study outcomes included lung function measures such as the post-bronchodilator forced expiratory volume in 1 second (FEV_1_) in liters (L) and the ratio of postbronchodilator FEV_1_/FVC. We also analyzed the respiratory quality of life quantified by the total St. George’s Respiratory Questionnaire (SGRQ) score (11). SGRQ score ranges between 0 and 100, where 0 indicates no symptom burden with a good quality of life, while 100 indicates a poor quality of patient life with high symptom burden (11). We also studied the association of fSAD^TLC^ with six-minute walk distance (6MWD) and mMRC (modified Medical Research Council) dyspnea scale that ranges from 0 to 4 in increasing order of shortness of breath (12).

In both cohorts, we further investigated the relationship between fSAD^TLC^ and change in FEV_1_ between baseline and follow-up visit after five years. The change in FEV_1_ was calculated as a difference between baseline FEV_1_ and the follow-up FEV_1_ around five years, which was then divided by the time between visits to calculate the change in mL / year.

### Statistical Analysis

To initially investigate the relationship between percent fSAD^TLC^ and fSAD^PRM^, we used Pearson’s correlation. Scatter plots were also generated to assess if fSAD^TLC^ was able to reliably capture the distribution of fSAD^PRM^. We also conducted a Bland-Altman analysis between the means of fSAD^TLC^ and fSAD^PRM^ to assess any systematic bias or variability in the differences across the range of measurements. Univariate and multivariable regression analysis was also conducted for investigating the association of fSAD^TLC^ with spirometry and clinical outcomes in COPD. We tested associations between baseline fSAD^TLC^ and post-bronchodilator FEV_1_ (L) and FEV_1_/FVC, with age, sex, race, body-mass index (BMI), smoking status, smoking pack years, and percent emphysema or LAA% as covariates. For assessing the association of baseline fSAD^TLC^ with SGRQ, 6MWD (ft), and mMRC dyspnea scale, we added baseline post-bronchodilator FEV_1_ (L) as an additional covariate for adjustment.

We studied the association of with change in FEV_1_ (mL / year) after adjusting for the same set of variables mentioned above. We repeated the multivariable analysis for change in FEV_1_ (mL / year) among GOLD 0 and GOLD 1 – 4 participants. To assess the relationship between fSAD^TLC^ and all-cause mortality, we categorized participants in four quartiles of fSAD^TLC^ and conducted Kaplan-Meier curve analysis for each quartile (13). Survival curves were plotted for each quartile, and the log-rank test was used to compare survival distributions across quartiles. All association studies and Kaplan-Meier curve analysis was repeated for the fSAD^PRM^. A two-sided *P* value < 0.05 was considered to be significant.

### Repeatability Analysis

We conducted a repeatability study to assess the overall reproducibility of fSAD^TLC^. We used data from the SPIROMICS repeatability study that enrolled 100 participants at the primary study sites between 2012 and 2015 (14). The study participants were scanned at TLC and RV during their enrollment visit. Repeat scans at both TLC and RV were acquired 2 – 6 weeks after the enrollment visit (14). For both visits, we generated synthetic RV scans for computing fSAD^TLC^. To test the repeatability of fSAD^TLC^, we computed the intraclass correlation coefficient (ICC) using the single measurement, same raters’ case (15). This was followed by a Bland-Altman analysis to test any systematic bias across the range of fSAD^TLC^. ICC and Bland-Altman plots were also generated for fSAD^PRM^ to compare its repeatability with fSAD^TLC^.

## RESULTS

### Study Design

SPIROMICS enrolled 2981 participants during its first phase from Nov 12, 2010, to July 31, 2015. Of 2981 participants, eight withdrew consent, and 1055 were reserved for training the TLC to RV generative model, as shown in **Figure E1**. Of 1918 scans, we removed 177 participants with change in TLC and RV volume less than 1L. After further eliminating the missing clinical information, we were able to obtain complete case data for 1458 participants at baseline in SPIROMICS (see **Figure E1**). At five-year follow-up, spirometry data was available only for 650 individuals (see **Figure E1**).

Of 10,305 individuals from the first phase of COPDGene, 473 had RV scans available for analysis at enrollment (see **Figure E1**). After eliminating the cases with missing clinical information, we analyzed baseline data from 458 participants to validate our results. For studying change in FEV_1_, eight subjects were lost to follow-up at five years, as shown in **Figure E1**.

### Participant Characteristics

The mean age at baseline was 62.9 (9.1) years in SPIROMICS and 63.3 (8.5) in COPDGene (see **Table 1**). Both cohorts were fairly balanced by biological sex with 788 (54%) and 215 (47%) males in SPIROMICS and COPDGene cohorts, respectively. In SPIROMICS, 588 (40%) individuals were current smokers, and the fraction was smaller for COPDGene with 130 (28%) individuals who smoked currently (see **Table 1**). As shown in **Table 1**, the mean baseline fSAD^TLC^ (%) in SPIROMICS was 12.98 (13.25) compared to a mean baseline fSAD^PRM^ (%) of 13.35 (12.21). Similarly, in COPDGene, the mean fSAD^TLC^ (%) was 9.55 (10.99) and the mean fSAD^PRM^ (%) was 10.13. (9.46) (see **Table 1**). Participant characteristics at five-year follow-up are reported in **Table E1** of the online supplement.

**Table 1:**
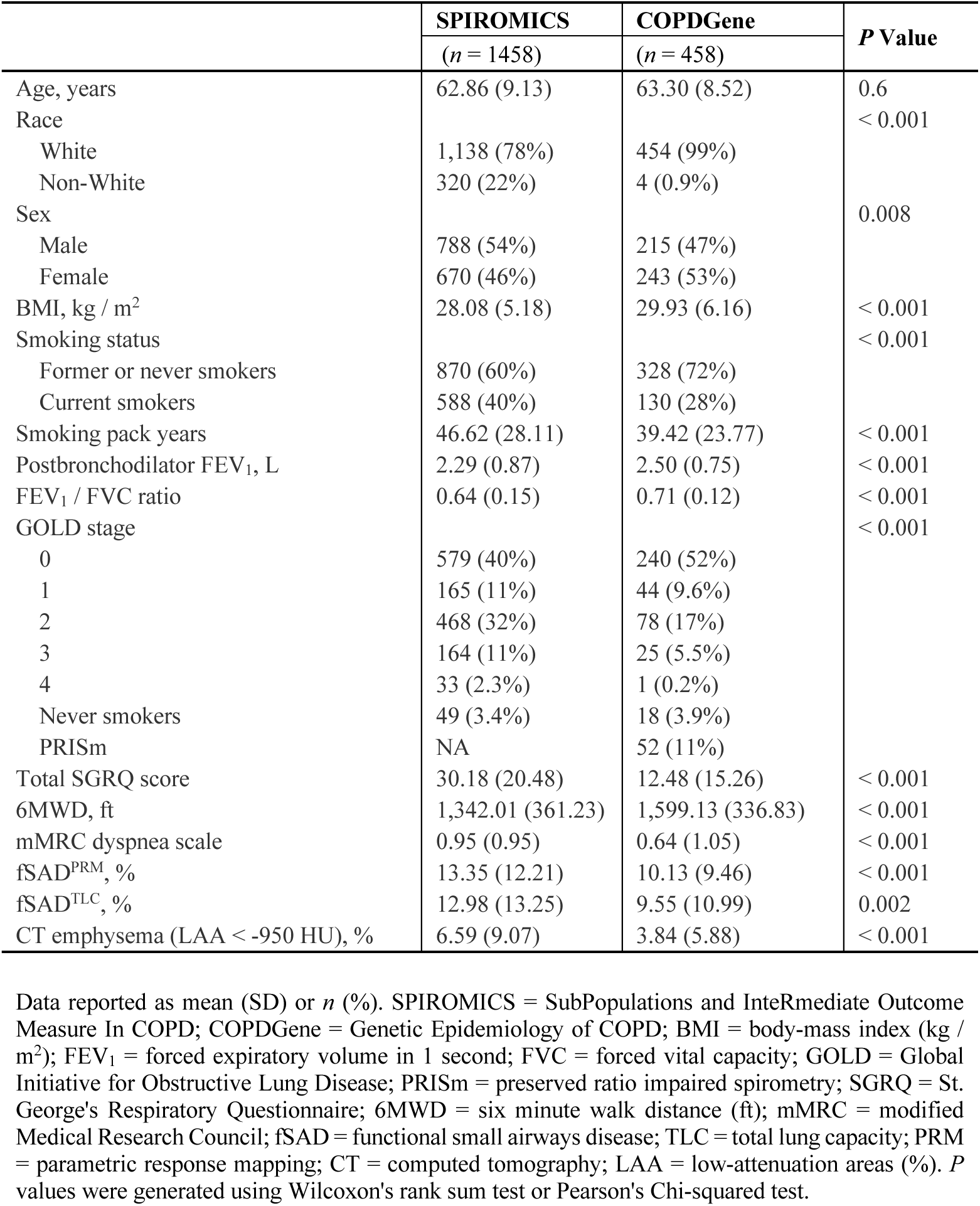
Participant characteristics from the SPIROMICS and COPDGene cohorts at baseline.

### Relationship between fSAD^TLC^ and fSAD^PRM^

We show a qualitative comparison between the spatial distributions of fSAD^PRM^ and fSAD^TLC^ (see columns 4 and 5 in **Figure 1**) across three different SPIROMICS subjects with varying degrees of fSAD. We also show mid-coronal slices from the virtual RV images generated by our AI model (see column 3 in **Figure 1**). Perceptually, there were negligible differences between the real and virtual RV image slices, shown respectively in columns 2 and 3 of **Figure 1**). We observed a high Pearson’s correlation between fSAD^TLC^ and fSAD^PRM^ in SPIROMICS (R = 0.895, *P* < 0.001) and COPDGene (R = 0.897, *P* < 0.001), suggesting a strong linear relationship between two variables (see **Figure 2A** and **2B**). Scatter plots showed a high agreement between the overall distributions of fSAD^TLC^ and fSAD^PRM^ in both cohorts (see **Figure 2A** and **2B**). Bland-Altman analysis between the means of fSAD^TLC^ and fSAD^PRM^ showed minimal bias of 0.37 and 0.58 in SPIROMICS (see **Figure 2C**) and COPDGene (see **Figure 2D**), respectively.

**Figure 1:**
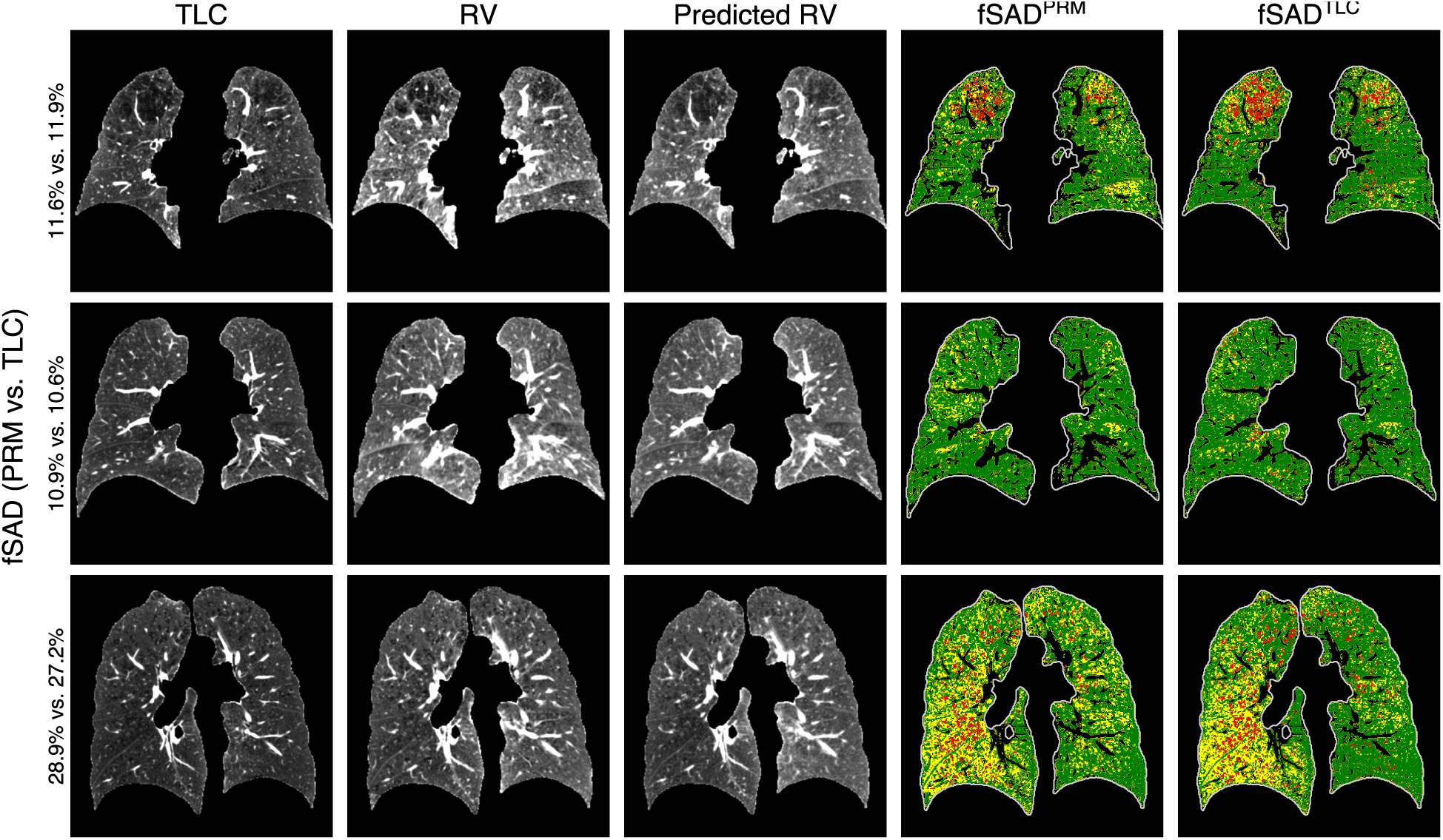
Spatial distribution of fSAD^PRM^ and fSAD^TLC^, shown on mid-coronal slices from three different individuals with varying degrees of small airways disease. The first and the second columns indicate chest CT scans at total lung capacity (TLC) and residual volume (RV), respectively. The TLC and RV scans were used to compute fSAD^PRM^. The third column shows the virtual or synthetic RV scans generated from the TLC chest CT scan alone. fSAD^TLC^ was computed using the TLC scan and the virtual RV scan, which allowed for a single-volume estimation of fSAD from TLC alone.

**Figure 2:**
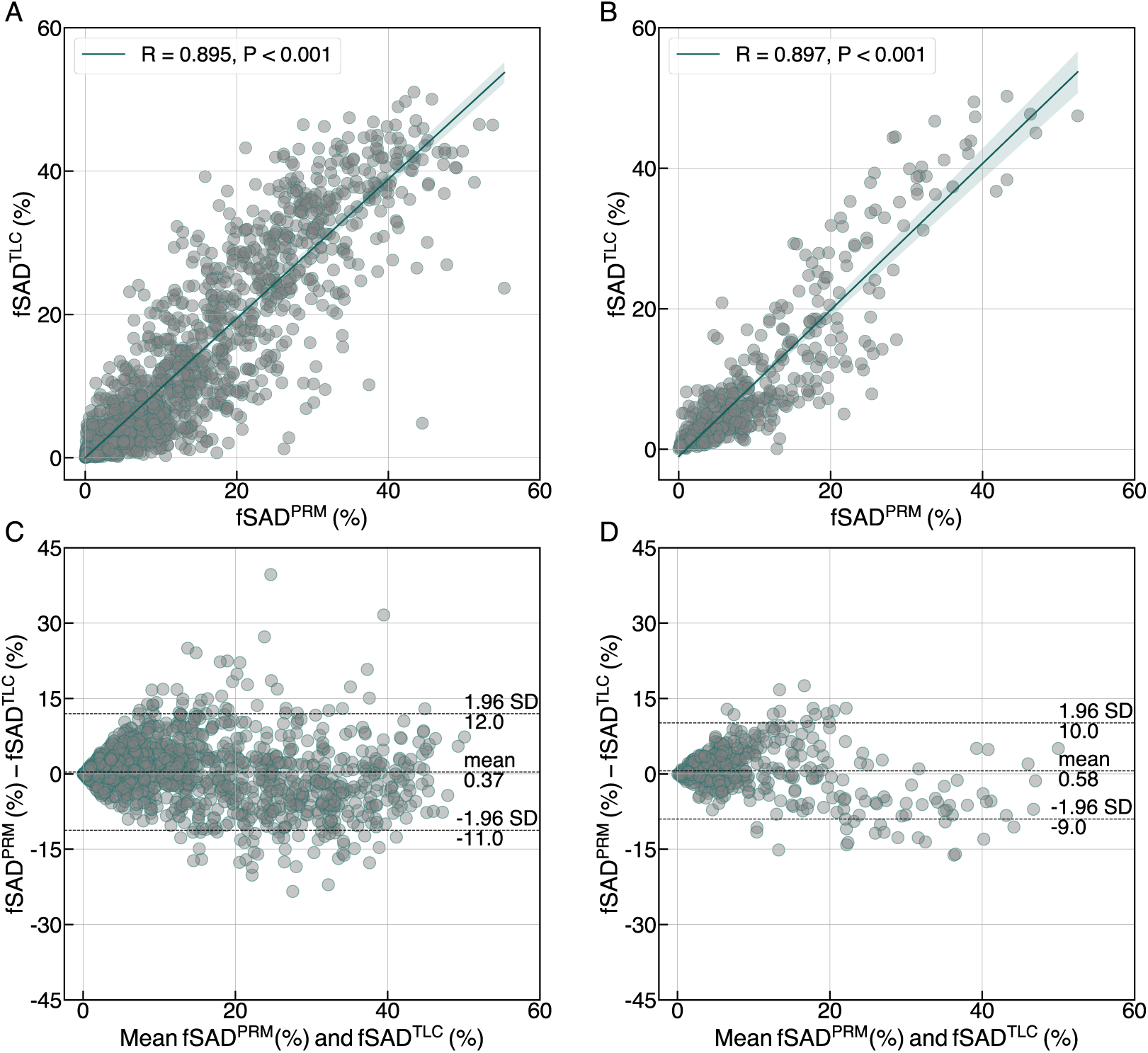
Relationship between fSAD^TLC^ with fSAD^PRM^ in both (A and C) SPIROMICS (*n* = 1458) and (B and D) COPDGene (*n* = 458) cohorts through scatter plots and Bland-Altman analysis. Pearson’s correlation, R is also reported for both cohorts.

### Lung Function and Respiratory Morbidity Predicted by fSAD^TLC^

Univariate regression analysis suggested significant association of fSAD^TLC^ with lung function and respiratory morbidity in SPIROMICS and COPDGene cohorts (see **Tables E2** and **E3**). On multivariable analysis, fSAD^TLC^ was significantly associated with lung function measures in SPIROMICS: postbronchodilator FEV_1_ (L) (adjusted β = −0.034, 95% CI: −0.037, −0.031; *P* < 0.001) and postbronchodilator FEV_1_/FVC (adjusted β = −0.008, 95% CI: −0.008, −0.007; *P* < 0.001) (see **Table 2**), independent of age, sex, race, BMI, smoking status, pack years, BMI, and percent emphysema. Similarly, fSAD^TLC^ was associated with FEV_1_ (L) (adjusted β = −0.032, 95% CI: - 0.038, −0.027; *P* < 0.001) and FEV_1_ / FVC (adjusted β = −0.007, 95% CI: −0.008, −0.007; *P* < 0.001) in COPDGene (see **Table 3**). fSAD^TLC^ was also associated with SGRQ in SPIROMICS (adjusted β = 0.240, 95% CI: 0.127, 0.353, *P* < 0.001) and COPDGene (adjusted β = 0.190, 95% CI: 0.030, 0.350, *P* = 0.02). To compare fSAD^TLC^ with fSAD^PRM^, we repeated the association studies for fSAD^PRM^ as well (see **Tables 2** and **3**). A Kaplan-Meier curve analysis revealed significantly increased rates of mortality (log rank *P* < 0.001) in individuals with increased fSAD^TLC^ (see **Figure 3**).

**Figure 3:**
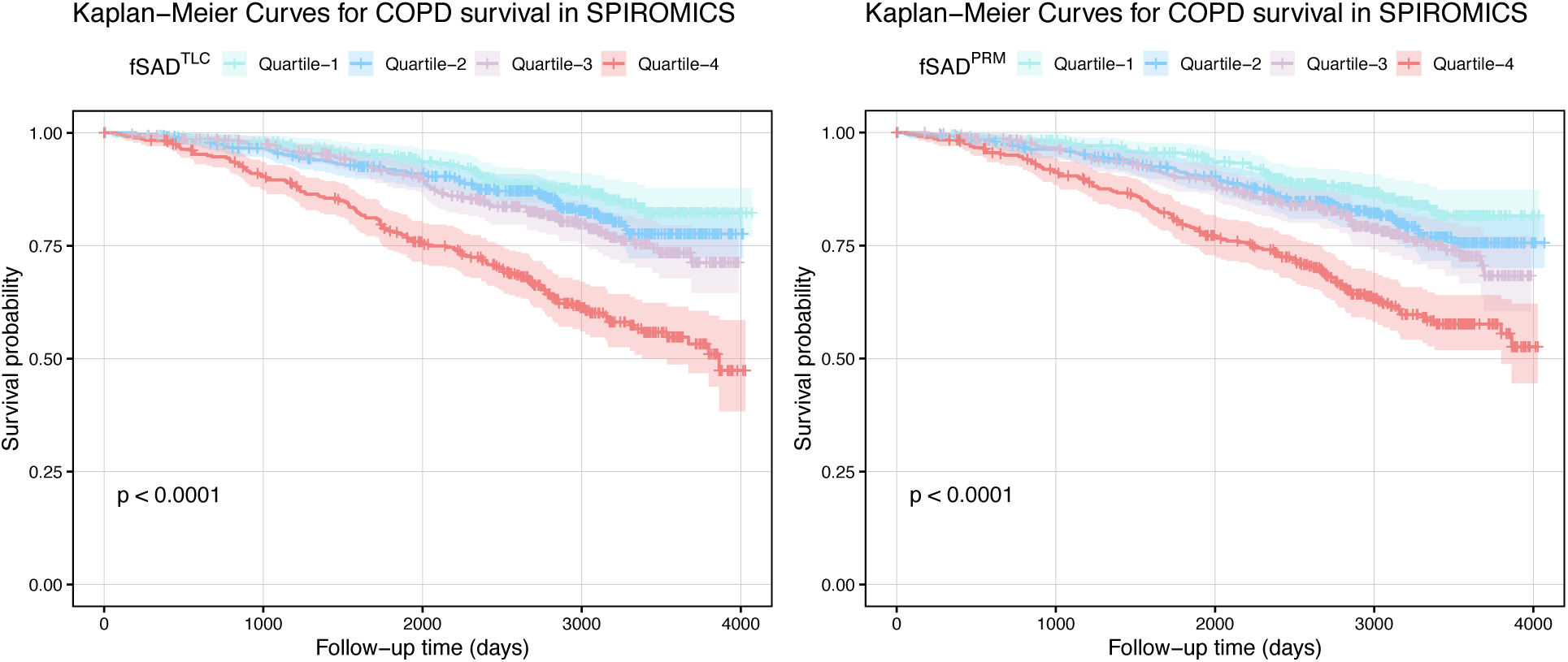
Kaplan-Meier curve analysis for studying overall survival in different quartiles of fSAD^TLC^ in both (A) SPIROMICS and (B) COPDGene cohorts.

**Table 2:** Multivariable linear regression analysis for assessing associations of baseline fSAD^TLC^ with lung function and respiratory morbidity in SPIROMICS (Estimate, 95% CI, *P* Value)

|  | fSAD<sup>TLC</sup> | *P* Value | fSAD<sup>PRM</sup> | *P* Value |
| --- | --- | --- | --- | --- |
| Postbronchodilator FEV<sub>1</sub>, L | -0.034 (-0.037, -0.031) | *P* < 0.001 | -0.034 (-0.037, -0.030) | *P* < 0.001 |
| Postbronchodilator FEV<sub>1</sub> / FVC | -0.008 (-0.008, -0.007) | *P* < 0.001 | -0.007 (-0.008, -0.007) | *P* < 0.001 |
| SGRQ | 0.240 (0.127, 0.353) | *P* < 0.001 | 0.177 (0.064, 0.290) | 0.002 |
| 6MWD, ft | -1.364 (-3.548, 0.819) | 0.22 | -1.887 (-4.057, 0.284) | 0.09 |
| mMRC Dyspnea Scale | 0.011 (-0.002, 0.024) | 0.10 | 0.01 (-0.003, 0.023) | 0.12 |

CI = confidence interval; fSAD = functional small airways disease; TLC = total lung capacity; FEV<sub>1</sub> = forced expiratory volume in 1 second; FVC = forced vital capacity; SGRQ = St. George's Respiratory Questionnaire; 6MWD = six-minute walk distance; mMRC = modified Medical Research Council. Multivariable models were adjusted for age, sex, race, smoking status, smoking pack years, body-mass index (BMI), baseline postbronchodilator FEV<sub>1</sub>, and percent emphysema define as percent LAA < -950 Hounsfield units (HU).

**Table 3:** Multivariable associations of baseline fSAD^TLC^ with lung function, symptom burden, and exercise capacity in the COPDGene cohort (Estimate, 95% CI, P Value)

|  | fSAD<sup>TLC</sup> | *P* Value | fSAD<sup>PRM</sup> | *P* Value |
| --- | --- | --- | --- | --- |
| Postbronchodilator FEV<sub>1</sub>, L | -0.032 (-0.038, -0.027) | *P* < 0.001 | -0.032 (-0.038, -0.026) | *P* < 0.001 |
| Postbronchodilator FEV<sub>1</sub> / FVC | -0.007 (-0.008, -0.007) | *P* < 0.001 | -0.007 (-0.008, -0.006) | *P* < 0.001 |
| SGRQ | 0.190 (0.030, 0.350) | 0.02 | 0.131 (-0.043, 0.304) | 0.14 |
| 6MWD, ft | -2.211 (-5.755, 1.332) | 0.22 | -2.505 (-6.332, 1.321) | 0.20 |
| mMRC Dyspnea Scale | 0.004 (-0.023, 0.03) | 0.76 | -0.008 (-0.037, 0.021) | 0.60 |

CI = confidence interval; fSAD = functional small airways disease; TLC = total lung capacity; FEV<sub>1</sub> = forced expiratory volume in 1 second; FVC = forced vital capacity; SGRQ = St. George's Respiratory Questionnaire; 6MWD = six-minute walk distance; mMRC = modified Medical Research Council. Multivariable models were adjusted for age, sex, race, smoking status, smoking pack years, body-mass index (BMI), baseline postbronchodilator FEV<sub>1</sub>, and percent emphysema define as percent LAA < -950 Hounsfield units (HU).

### Change in FEV_1_ and fSAD^TLC^

In both SPIROMICS (β = −1.156, 95% CI: −1.699, −0.613; *P* < 0.001) and COPDGene (β = −0.866, 95% CI: −1.386, −0.345; *P* < 0.001), fSAD^TLC^ was significantly associated with change in FEV_1_ (mL / year) for all subjects (see **Table 4**). We conducted stratified linear regression analysis for GOLD 0 and GOLD 1 – 4 to investigate the impact of fSAD^TLC^ on FEV_1_ decline based on baseline airflow limitation (see **Table 4**). In both cohorts, fSAD^TLC^ was associated with FEV_1_ decline for GOLD 1 – 4 participants (see **Table 4**). In SPIROMICS, for every additional 1% increase in fSAD^TLC^, a decline in FEV_1_ of 1.050 mL / year (*P* = 0.001) was observed for GOLD 1 – 4 participants. Similarly, in COPDGene, for a 1% increase fSAD^TLC^, FEV_1_ declined by 1.175 mL / year (*P* = 0.003) (see **Table 4**).

**Table 4:** Association between fSAD^TLC^ and change in FEV_1_ (mL / year) stratified by baseline COPD GOLD stage (Estimate, 95% CI, *P* Value)

|  |  | fSAD<sup>TLC</sup> | *P* Value | fSAD<sup>PRM</sup> | *P* Value |
| --- | --- | --- | --- | --- | --- |
| **SPIROMICS** | **Overall** | -1.156 (-1.699, -0.613) | *P* < 0.001 | -0.742 (-1.280, -0.205) | 0.007 |
|  | **GOLD 0** | -2.994 (-5.900, -0.088) | 0.04 | -0.084 (-1.244, 1.077) | 0.89 |
|  | **GOLD 1 - 4** | -1.050 (-1.691, -0.408) | 0.001 | -0.899 (-1.598, -0.201) | 0.01 |
| **COPDGene** | **Overall** | -0.866 (-1.386, -0.345) | 0.001 | -0.871 (-1.440, -0.303) | 0.003 |
|  | **GOLD 0** | -0.230 (-1.760, 1.300) | 0.77 | -0.514 (-1.845, 0.817) | 0.45 |
|  | **GOLD 1 - 4** | -1.175 (-1.943, -0.406) | 0.003 | -1.067 (-1.934, -0.200) | 0.02 |

CI = confidence interval; fSAD = functional small airways disease; TLC = total lung capacity; GOLD = Global Initiative for Obstructive Lung Disease; FEV<sub>1</sub> = forced expiratory volume in 1 second. Multivariable models were adjusted for age, sex, race, smoking status, smoking pack years, body-mass index (BMI), baseline postbronchodilator FEV<sub>1</sub>, and percent emphysema define as percent LAA < -950 Hounsfield units (HU).

### Repeatability of fSAD^TLC^

Over a short follow-up of 2 – 6 weeks, we observed fSAD^TLC^ to be highly repeatable with ICC of 0.99 (95% CI: 0.98, 0.99). This was significantly higher than the ICC of fSAD^PRM^ (0.83 (95% CI: 0.76, 0.88)). A Bland-Altman analysis between the fSAD^TLC^ and fSAD^PRM^ computed at visit 1 and visit 2 chest CT scans from the SPIROMICS Repeatability study revealed minimal bias for both fSAD^PRM^ (0.26) and fSAD^TLC^ (0.22) (see **Figure 4A** and **4B**). The limits of agreement were significantly larger for fSAD^PRM^ (18%) as compared to fSAD^TLC^ (5%), suggesting greater overall variability in fSAD^PRM^ between 2 – 6 weeks follow-up.

**Figure 4:**
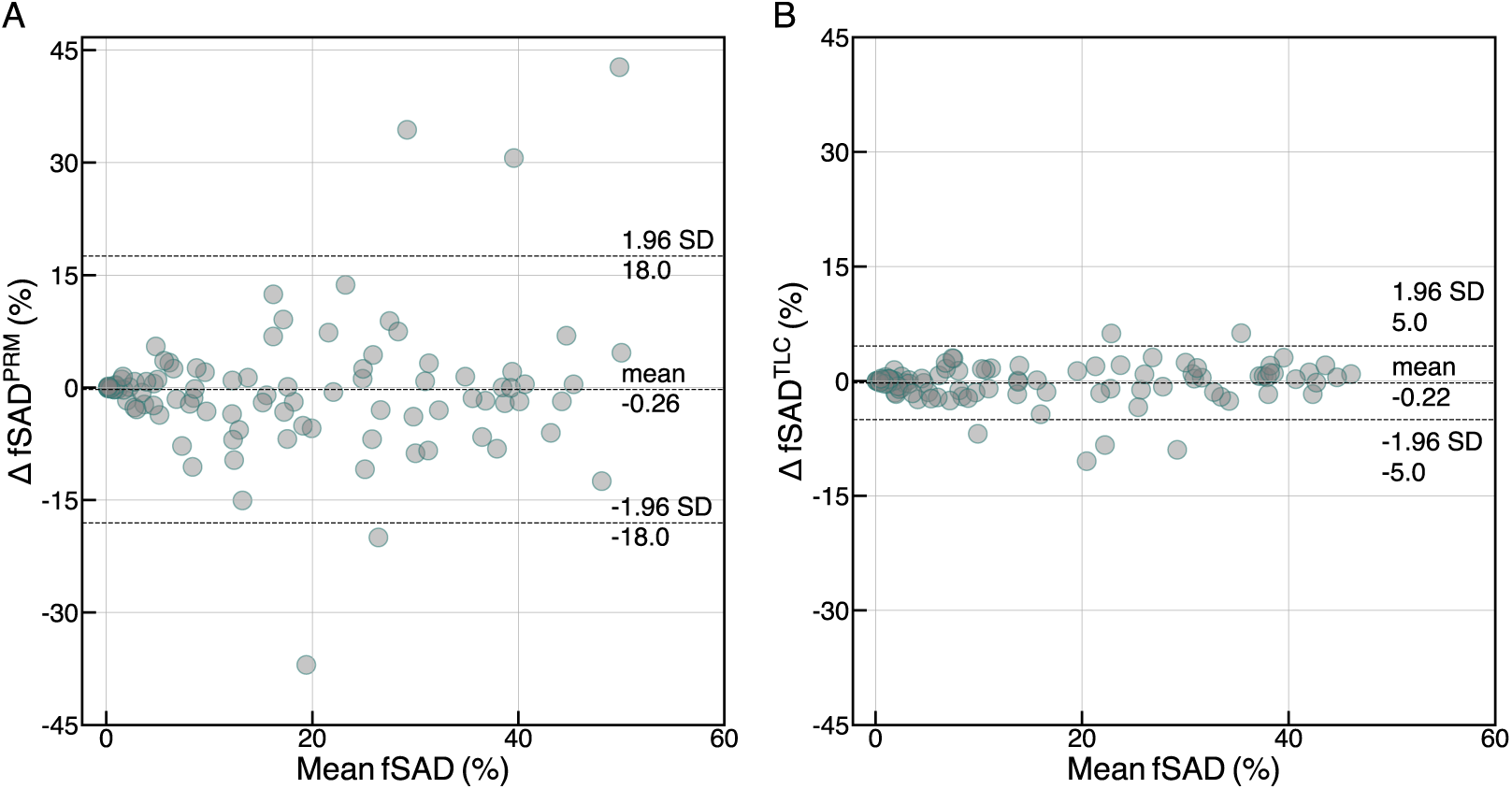
Repeatability study of (A) fSAD^PRM^ and (B) fSAD^TLC^ between baseline and 2 to- 6- week follow-up (*n* = 98) through Bland-Altman analysis.

## Discussion

We aimed to determine whether generative AI can be used to derive fSAD from a single CT scan at TLC, and whether this single volume fSAD estimation correlates with clinical and functional outcomes in COPD. Traditionally, fSAD estimation has relied on an additional expiratory CT scan, a requirement that limits its clinical applicability (4, 10). We believe this is the first study to utilize a generative AI model for estimating fSAD from a single inspiratory CT scan. We demonstrated that, in two large cohorts, fSAD measured by TLC CT alone is associated with poor lung function, respiratory quality of life, and FEV_1_ decline.

A comparison of fSAD^TLC^ and fSAD^PRM^ suggested a strong linear relationship between the two measurements, supporting our hypothesis that SAD can be accurately measured from TLC CT using generative AI. During training, our model learned to transform TLC scans to RV scans from a large number of TLC and RV pairs. Exposing our model to a large training dataset improved virtual RV image generation, allowing us to capture SAD from TLC as accurately as fSAD^PRM^. An equally strong relationship between fSAD^TLC^ and fSAD^PRM^ was observed in the COPDGene cohort suggested model generalizability to different cohort. For both cohorts, Bland-Altman analysis between the means of percent fSAD^TLC^ and fSAD^PRM^ showed minimal bias across the range of two measurements, and the limits of agreement, while not too narrow, were reasonable, suggesting that the two methods are generally consistent and could potentially be used interchangeably within an acceptable range.

We demonstrated that fSAD^TLC^ was significantly associated with FEV_1_ decline in both SPIROMICS and COPDGene cohorts. Our findings were consistent with those of Bhatt *et al*., who found similar associations of fSAD^PRM^ with FEV_1_ decline in COPDGene (16). Further, in individuals with established airflow limitation and mild-to-severe COPD (GOLD 1 – 4), fSAD^TLC^ was strongly associated with FEV_1_ decline independent of baseline percent emphysema. This suggested that there was an involvement of a small airways disease component at baseline contributing towards FEV_1_ decline. A consistency of these findings with fSAD^PRM^ indicated the potential of single chest CT volume fSAD^TLC^ as an alternative for characterizing disease progression in COPD. For both univariate and multivariable models, the direction of regression coefficients were consistent, and magnitudes were comparable between fSAD^TLC^ and fSAD^PRM^. This suggested a consistent influence of unit change in fSAD^TLC^ and fSAD^PRM^ on the outcome variables further signifying an agreement between them.

An important part of this study was the repeatability analysis conducted using data short-term follow-up data from the SPIROMICS Repeatability study (14). The repeat scans over short follow-up prevented any long-term variabilities that might occur due to disease. fSAD^TLC^ was significantly more repeatable than fSAD^PRM^, however, this was expected due to the potential for variations in patient effort between the repeat RV scans. Less common acquisition of RV scans in clinical practice results in inadequate coaching to expiratory volumes (RV and FRC), making them hard to reproduce. Since fSAD^PRM^ relies on RV scans, its repeatability is more dependent on adequate patient effort each time they are being scanned. Since expiration scans are thus harder to reproduce (14), this complicates the repeatability of fSAD^PRM^. On the contrary, the proposed fSAD^TLC^ relies only on inspiratory chest CT scans, which are more easily reproducible and less prone to inadequate patient effort (17). These findings were also consistent with the results from the Repeatability Study where the dual volume CT biomarkers showed significantly lower ICC values compared to single volume metrics (14).

A few limitations of this study need to be recognized. Our spatial estimation of fSAD^TLC^ consistent with fSAD^PRM^ with a few differences in some isolated regions. Still, the major fSAD clusters were picked up by our method. We developed our model using scans from the SPIROMICS study that were acquired using a quality-controlled imaging protocol. This may not be true for the TLC scans acquired in clinical settings. It would be important to test our method in TLC scans with lower dosage and different acquisition protocols. We had to remove subjects with unreliable TLC or RV (volume difference < 1L) and acknowledge this as a quality control step in our analysis; fortunately, only 10% of the scans were considered unusable. For assessing change in FEV_1_, we were limited to only two lung function measurements separated by a follow-up of almost five years. Another limitation of our method was that all the scans were acquired without any contrast agent, and whether these results are reproducible in CT scans with contrast remains to be evaluated.

As our findings suggest, the current study has a number of strengths. Inspiratory CT scans are common in clinical settings, and our method allows for the detection of functional small airway disease by using them to create virtual expiratory scans. Large-scale characterization and phenotyping of individuals with small airways disease has been limited due to the fact that fSAD^PRM^ requires RV scans which are not routinely acquired in most settings. Also, retrospective evaluation of individuals with a TLC scan but no expiratory scan is not possible. The proposed method addresses these concerns by allowing an assessment of fSAD in patient cohorts with only TLC scans, an example being the National Lung Screening Trial (NLST) (19). Our method can also be used for the retrospective evaluation of large patient cohorts where expiratory chest CT scans were not acquired, an instance being the Multi-Ethnic Study of Atherosclerosis (MESA) (18). Also, this method has the potential to reduce radiation exposure to patients in future studies of small airway disease.

## Supporting information

Supplementary Methods and Results

## Data Availability

All data produced in the present study are available upon reasonable request to the authors.

## Acknowledgements

The authors thank the SPIROMICS participants and participating physicians, investigators, and staff for making this research possible. More information about the study and how to access SPIROMICS data is available at www.spiromics.org. The authors would like to acknowledge the University of North Carolina at Chapel Hill BioSpecimen Processing Facility for sample processing, storage, and sample disbursements (**http://bsp.web.unc.edu/**).

We would like to acknowledge the following current and former investigators of the SPIROMICS sites and reading centers: Neil E Alexis, MD; Wayne H Anderson, PhD; Mehrdad Arjomandi, MD; Igor Barjaktarevic, MD, PhD; R Graham Barr, MD, DrPH; Patricia Basta, PhD; Lori A Bateman, MSc; Surya P Bhatt, MD; Eugene R Bleecker, MD; Richard C Boucher, MD; Russell P Bowler, MD, PhD; Stephanie A Christenson, MD; Alejandro P Comellas, MD; Christopher B Cooper, MD, PhD; David J Couper, PhD; Gerard J Criner, MD; Ronald G Crystal, MD; Jeffrey L Curtis, MD; Claire M Doerschuk, MD; Mark T Dransfield, MD; Brad Drummond, MD; Christine M Freeman, PhD; Craig Galban, PhD; MeiLan K Han, MD, MS; Nadia N Hansel, MD, MPH; Annette T Hastie, PhD; Eric A Hoffman, PhD; Yvonne Huang, MD; Robert J Kaner, MD; Richard E Kanner, MD; Eric C Kleerup, MD; Jerry A Krishnan, MD, PhD; Lisa M LaVange, PhD; Stephen C Lazarus, MD; Fernando J Martinez, MD, MS; Deborah A Meyers, PhD; Wendy C Moore, MD; John D Newell Jr, MD; Robert Paine, III, MD; Laura Paulin, MD, MHS; Stephen P Peters, MD, PhD; Cheryl Pirozzi, MD; Nirupama Putcha, MD, MHS; Elizabeth C Oelsner, MD, MPH; Wanda K O’Neal, PhD; Victor E Ortega, MD, PhD; Sanjeev Raman, MBBS, MD; Stephen I. Rennard, MD; Donald P Tashkin, MD; J Michael Wells, MD; Robert A Wise, MD; and Prescott G Woodruff, MD, MPH. The project officers from the Lung Division of the National Heart, Lung, and Blood Institute were Lisa Postow, PhD, and Lisa Viviano, BSN; SPIROMICS was supported by contracts from the NIH/NHLBI (HHSN268200900013C, HHSN268200900014C, HHSN268200900015C, HHSN268200900016C, HHSN268200900017C, HHSN268200900018C, HHSN26820-0900019C, HHSN268200900020C), grants from the NIH/NHLBI (U01 HL137880 and U24 HL141762), and supplemented by contributions made through the Foundation for the NIH and the COPD Foundation from AstraZeneca/MedImmune; Bayer; Bellerophon Therapeutics; Boehringer-Ingelheim Pharmaceuticals, Inc.; Chiesi Farmaceutici S.p.A.; Forest Research Institute, Inc.; GlaxoSmithKline; Grifols Therapeutics, Inc.; Ikaria, Inc.; Novartis Pharmaceuticals Corporation; Nycomed GmbH; ProterixBio; Regeneron Pharmaceuticals, Inc.; Sanofi; Sunovion; Takeda Pharmaceutical Company; and Theravance Biopharma and Mylan.

This work was supported by NHLBI grants U01 HL089897 and U01 HL089856 and by NIH contract 75N92023D00011. The COPDGene study (NCT00608764) has also been supported by the COPD Foundation through contributions made to an Industry Advisory Committee that has included AstraZeneca, Bayer Pharmaceuticals, Boehringer-Ingelheim, Genentech, GlaxoSmithKline, Novartis, Pfizer, and Sunovion.

