## Supplementary Methods and Results for "Deep Learning Estimation of Small Airways Disease from Inspiratory Chest CT is Associated with FEV_1_ Decline in COPD"

[ϒ]Muhammad F. A. Chaudhary is now at the Center for Lung Analytics and Imaging Research (CLAIR) at The University of Alabama at Birmingham. This study was conducted when he was at The Roy J. Carver Department of Biomedical Engineering at The University of Iowa.

### Supplementary Methods

#### Data Preparation and Preprocessing

We analyzed and processed the chest CT scans from the SubPopulations and InteRmediate Outcome Measures in COPD Study (SPIROMICS) to train our generative model (1). Originally, images were acquired at a resolution of approximately $0.6 \times 0.6 \times 0.5$ mm$^3$. We first registered the chest CT scans at residual volume (RV) to the CT scans at total lung capacity (TLC) using a B-Spline parameterized deformable image registration method. We used a mass-preserving cost function to account for the intensity change between the inspiratory and expiratory chest CT scans. After registration, both TLC and RV scans were isotropically resampled to 1mm$^3$, clipped between a Hounsfield unit (HU) range of -1024 HU and 1024 HU, and normalized between -1 and 1. For more details, please refer to (2).

#### Generative Adversarial Networks (GANs) and LungViT

Generative Adversarial Networks (GANs) constitute a class of deep learning frameworks that can be used to generate images of different styles and texture given a template input image (2, 3). To do so, GANs require a paired training dataset that can be used to learn a transformation from on image to another. Our goal was to transform a chest CT scan at inspiration to a chest CT scan at expiration, so that it could be used to compute functional small airways disease (fSAD) from a chest CT at inspiration *alone*.

GANs comprise two primary components or neural networks: the generator and the discriminator. The generator creates RV images from the given TLC images, while the discriminator evaluates the authenticity and perceptual realism of the generated RV samples. Both the generator and discriminator are trained simultaneously through adversarial training, where the generator endeavors to deceive the discriminator with realistic data, while the discriminator aims to correctly identify real and fake data. Over time, this competitive process assists the generator in producing increasingly realistic data (3).

In this study, we use a specialized GAN framework developed specifically for transforming an inspiratory chest CT at total lung capacity (TLC) to an expiratory chest CT at residual volume (RV). The model was recently published, and it used novel deep learning architectures and loss functions to generate high fidelity RV scans from the given TLC CT scans. For more details on model training strategies, network architectures, and loss functions, please refer to our study in (2).

### Supplementary Results

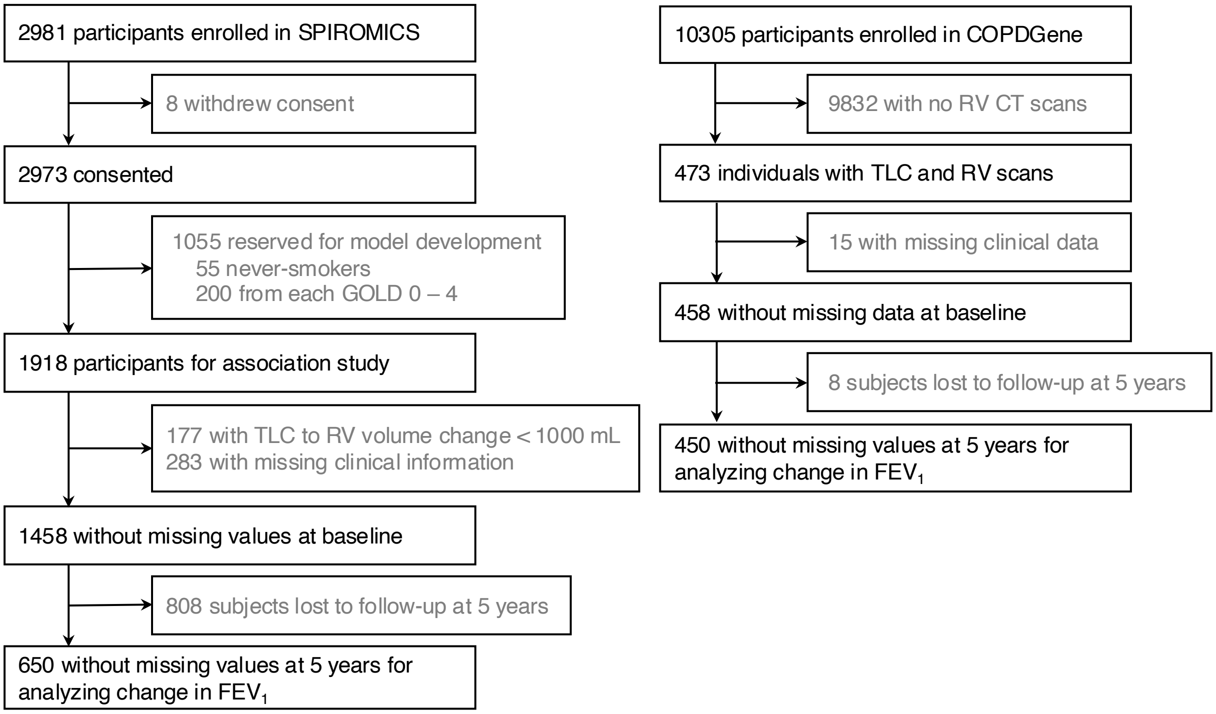


**Figure E1**: CONSORT diagram. A subset of SPIROMICS cohort (*n* = 1055) was used for model development, which was applied to the SPIROMICS (remaining *n* = 1460) and COPDGene (n = 458) cohorts for investigating the association of $fSAD^{TLC}$ with clinical outcomes in COPD.

**Table E1**: Participant characteristics from the SPIROMICS and COPDGene cohorts at five-year follow-up for assessing change in $FEV_1$.

| | **SPIROMICS** | **COPDGene** | ***P* Value** |
|---|---|---|---|
| | (*n* = 650) | (*n* = 450) | |
| Age, years | 63.28 (8.54) | 62.99 (8.85) | > 0.9 |
| Race | | | < 0.001 |
| White | 447 (99%) | 497 (76%) | |
| Non-White | 3 (0.7%) | 153 (24%) | |
| Sex | | | 0.15 |
| Male | 212 (47%) | 335 (52%) | |
| Female | 238 (53%) | 315 (48%) | |
| BMI, kg / $m^2$ | 29.90 (6.05) | 28.46 (5.00) | 0.002 |
| Smoking status | | | 0.011 |
| Former or never smokers | 325 (72%) | 422 (65%) | |
| Current smokers | 125 (28%) | 228 (35%) | |
| Smoking pack years | 39.46 (23.88) | 46.14 (29.19) | < 0.001 |
| Postbronchodilator $FEV_1$, L | 2.50 (0.75) | 2.36 (0.82) | 0.002 |
| $FEV_1$ / FVC ratio | 0.71 (0.12) | 0.66 (0.14) | < 0.001 |
| GOLD stage | | | |
| 0 | 238 (53%) | 271 (42%) | |
| 1 | 43 (9.6%) | 89 (14%) | |
| 2 | 74 (16%) | 203 (31%) | |
| 3 | 24 (5.3%) | 54 (8.3%) | |
| 4 | 1 (0.2%) | 9 (1.4%) | |
| Never smokers | 70 (16%) | 24 (3.7%) | |
| Total SGRQ score | 12.33 (15.18) | 27.52 (19.25) | < 0.001 |
| 6MWD, ft | 1,600.96 (335.91) | 1,392.87 (329.52) | < 0.001 |
| mMRC dyspnea scale | 0.95 (0.95) | 0.64 (1.05) | < 0.001 |
| $fSAD^{PRM}$, % | 10.01 (9.33) | 12.02 (11.16) | 0.065 |
| $fSAD^{TLC}$, % | 9.42 (10.91) | 11.40 (12.27) | 0.3 |
| CT emphysema (LAA < -950 HU), % | 3.84 (5.92) | 5.56 (7.98) | 0.003 |
| Change in $FEV_1$ (mL / year) | -35.29 (40.29) | -32.77 (51.65) | 0.3 |

Data reported as mean (SD) or *n* (%). SPIROMICS = SubPopulations and InteRmediate Outcome Measure In COPD; COPDGene = Genetic Epidemiology of COPD; BMI = body-mass index (kg / $m^2$); $FEV_1$ = forced expiratory volume in 1 second; FVC = forced vital capacity; GOLD = Global Initiative for Obstructive Lung Disease; PRISm = preserved ratio impaired spirometry; SGRQ = St. George's Respiratory Questionnaire; 6MWD = six minute walk distance (ft); mMRC = modified Medical Research Council; fSAD = functional small airways disease; TLC = total lung capacity; PRM = parametric response mapping; CT = computed tomography; LAA = low-attenuation areas (%). *P* values were generated using Wilcoxon's rank sum test or Pearson's Chi-squared test.

**Table E2**: Univariate linear regression analysis for assessing associations of baseline $fSAD^{TLC}$ with lung function and respiratory morbidity in SPIROMICS (Estimate, 95% CI, *P* Value).

| | $fSAD^{TLC}$ | *P* Value | $fSAD^{PRM}$ | *P* Value |
|---|---|---|---|---|
| Postbronchodilator $FEV_1$, L | -0.040 (-0.043, -0.038) | $P < 0.001$ | -0.042 (-0.045, -0.039) | $P < 0.001$ |
| Postbronchodilator $FEV_1$ / FVC | -0.010 (-0.010, -0.009) | $P < 0.001$ | -0.010 (-0.010, -0.010) | $P < 0.001$ |
| SGRQ | 0.484 (0.408, 0.559) | $P < 0.001$ | 0.442 (0.359, 0.525) | $P < 0.001$ |
| 6MWD, ft | -6.857 (-8.213, -5.501) | $P < 0.001$ | -7.049 (-8.526, -5.572) | $P < 0.001$ |
| mMRC Dyspnea Scale | 0.039 (0.032, 0.047) | $P < 0.001$ | 0.038 (0.03, 0.046) | $P < 0.001$ |

CI = confidence interval; fSAD = functional small airways disease; TLC = total lung capacity; $FEV_1$ = forced expiratory volume in 1 second; FVC = forced vital capacity; SGRQ = St. George's Respiratory Questionnaire; 6MWD = six-minute walk distance; mMRC = modified Medical Research Council.

**Table E3**: Univariate linear regression analysis for assessing associations of baseline $fSAD^{TLC}$ with lung function and respiratory morbidity in COPDGene (Estimate, 95% CI, *P* Value).

| | $fSAD^{TLC}$ | *P* Value | $fSAD^{PRM}$ | *P* Value |
|---|---|---|---|---|
| Postbronchodilator $FEV_1$, L | -0.031 (-0.037, -0.026) | $P < 0.001$ | -0.037 (-0.043, -0.030) | $P < 0.001$ |
| Postbronchodilator $FEV_1$ / FVC | -0.009 (-0.009, -0.008) | $P < 0.001$ | -0.009 (-0.010, -0.009) | $P < 0.001$ |
| SGRQ | 0.520 (0.401, 0.638) | $P < 0.001$ | 0.500 (0.359, 0.641) | $P < 0.001$ |
| 6MWD, ft | -5.910 (-8.677, -3.143) | $P < 0.001$ | -7.336 (-10.543, -4.129) | $P < 0.001$ |
| mMRC Dyspnea Scale | 0.049 (0.033, 0.065) | $P < 0.001$ | 0.047 (0.028, 0.066) | $P < 0.001$ |

CI = confidence interval; fSAD = functional small airways disease; TLC = total lung capacity; $FEV_1$ = forced expiratory volume in 1 second; FVC = forced vital capacity; SGRQ = St. George's Respiratory Questionnaire; 6MWD = six-minute walk distance; mMRC = modified Medical Research Council.
